# Total testosterone is not associated with lean mass or handgrip strength in pre-menopausal females: findings from the NHANES

**DOI:** 10.1101/2020.08.04.20168542

**Authors:** Sarah E. Alexander, Gavin Abbott, Brad Aisbett, Glenn D. Wadley, Jill A. Hnatiuk, Séverine Lamon

**Author notes:** Corresponding Author: Dr Séverine Lamon, PhD; Institute for Physical Activity and Nutrition (IPAN), School of Exercise and Nutrition Sciences, Deakin University, 221 Burwood Hwy, Burwood 3125, Australia. ph. Authors contributed equally (Jill A. Hnatiuk, Séverine Lamon).

## Abstract

Testosterone is a naturally occurring hormone that has been positively associated with lean mass and strength in males. Whether endogenous testosterone is related to lean mass and strength in females is unknown.

**Objective:** To examine the relationship between endogenous testosterone concentration and lean mass and handgrip strength in healthy, pre-menopausal females.

**Methods:** Secondary data from the 2013-2014 National Health and Nutrition Examination Survey (NHANES)were used. Females were aged 18-40 (n=753, age 30 ± 6 yr, mean ± SD) and pre-menopausal. Multivariate linear regression models were used to examine associations between total testosterone, height-adjusted lean mass and handgrip strength.

**Results:** Mean ± SD testosterone concentration was 1.0 ± 0.6 nmol·L^-1^ and mean free androgen index (FAI) was 0.02 ± 0.02. Mean fat-free mass index (FFMI) was 16.4 ± 3.0 kg·m^-2^ and mean handgrip strength was 61.7 ± 10.5 kg. In females, testosterone was not associated with FFMI (β=0.08; 95%CI: −0.02, 0.18; *p=0.11)* or handgrip strength (β=0.03; 95%CI: −0.11, 0.17; *p=0.67)* in a statistically significant manner. Conversely, FAI was positively associated with FFMI (β=0.17; 95%CI: 0.01, 0.33; *p=0.04*) but not handgrip strength (β=0.19; 95%CI: −0.02, 0.21; *p=0.10*).

**Conclusions:** These findings indicate that FAI, but not total testosterone, is associated with FFMI in females. The small coefficients however suggest that FAI only accounts for a minor proportion of the variance in FFMI, highlighting the complexity of the regulation of lean mass in female physiology. FAI nor total testosterone are associated with handgrip strength in females when testosterone concentrations are not altered pharmacologically.

## Introduction

The maintenance of skeletal muscle mass and function is not only essential for health and quality of life across the lifespan, it is also a determining factor of athletic performance (1). Despite females representing 50% of the human population, research in the field of skeletal muscle regulation and the response to exercise has been overwhelmingly performed on male cohorts. Between 2017 and 2019, only 8% of all sports and exercise research was made up of female-only cohorts and the majority of these tend to relate to aspects specific to females, such as pregnancy, menopause or reproductive disease (2). However, male and female muscle physiology differ in many ways. For example, the growth and regenerative capacity of skeletal muscle vary between males and females (3). Male myocytes exhibit greater proliferative capacity, while female myocytes display greater differentiation *in vitro* (3). There are also sex-specific differences in skeletal muscle morphology, where females have more type I muscle fibres, while males have more type IIb muscle fibres (4). In response to resistance training, females display greater fatigue resistance and a greater capacity for neural adaptations when compared to males (5). These differences are driven, in part, by varying concentrations of the major sex hormones, oestrogen and testosterone (3, 4).

The major androgen hormone testosterone is an anabolic hormone that regulates skeletal muscle growth. It exerts its effects on target tissues, including skeletal muscle, by binding to its specific receptor, the androgen receptor (AR) (6). Testosterone is also present in females, albeit at concentrations about 10-fold lower than typical male levels (7). In females, testosterone is mostly active in the regulation of the reproductive and nervous systems (7); however, its role in the regulation of female skeletal muscle growth is not well understood. Despite having about 10-fold less testosterone than males, females exhibit similar relative strength (8) and muscle mass gains (9) as males in response to resistance training. Protein synthesis and degradation rates are also similar between males and females, both at rest and after resistance exercise (10). There is further evidence from mouse studies to suggest that testosterone and other androgen hormones may not be necessary to reach peak muscle mass or strength in females (11). Instead, growth hormone (GH), insulin-like growth factor-1 (IGF-1) and oestrogen may take over some of the anabolic role of testosterone in females (11, 12).

In untrained males, a moderate-to-strong positive relationship exists between testosterone concentrations, lean body mass and muscle strength, when expressed relative to body mass (13, 14). In young healthy men (n=61), testosterone concentrations correlated with fat-free mass, leg muscle size and strength in a dose-dependent manner when testosterone concentrations were pharmaceutically manipulated for 20 weeks (14). This holds true for endogenous testosterone, where men with high testosterone concentrations have more relative lean mass than those with low testosterone concentrations (n=252) (13). Limited evidence about the relationship between testosterone, muscle mass and muscle strength is currently available in females. Administration of exogenous testosterone that raised testosterone levels by approximately four-fold for 10 weeks resulted in increased lean mass and running time to exhaustion (15), but did not alter body fat percentage, VO2max or functional outcomes including leg muscle strength and power and anaerobic power (15). The relationship between endogenous testosterone and muscle-related outcomes has not been investigated using large cohorts of healthy females using appropriately adjusted models.

The aim of this study was to examine cross-sectional evidence of relationships between endogenous testosterone concentrations, lean mass and handgrip strength in 18-40-year-old premenopausal females from the National Health and Nutrition Examination Survey (NHANES). It was hypothesised that there would be no associations between total testosterone and lean mass or handgrip strength in this population.

## Methods

### Study Population

The Deakin University Human Research Ethics Committee (DUHREC) exempted this study from ethics review. Information regarding the consent process of the NHANES is located at https://www.cdc.gov/nchs/nhanes.

NHANES is a nationally representative, cross-sectional survey conducted in the United States of America that has run annually since 1971. NHANES uses a multi-stage, stratified, clustered probability sample including non-institutionalised civilians over two months of age. Further information about sampling, study design and all protocols can be found at https://www.cdc.gov/nchs/nhanes. Briefly, NHANES consists of an initial at-home interview, where trained staff ask questions with automated data collection. All participants then attend a mobile examination clinic (MEC) where trained staff collect anthropometric data and biological samples. This study used the cohort recruited in 2013-2014, where 10,175 individuals participated in the at-home interviews. Of these individuals, 9,813 participated in the MEC (96%).

Individuals were excluded from the cohort if they were male (n=5,003) or if they were younger than 18 (n=1,975) or older than 40 (n=1,949) years of age. This age range includes young to middle-aged females that were not menopausal, as menopause may affect the relationship between testosterone and skeletal muscle due to the significant decrease in oestrogen and sex-hormone binding globulin (SHBG) that occurs at this time (16). Females who were pregnant (n=63), or who had not had regular menses in the last 12 months due to menopause (n=1), responses were excluded. Individuals with previous diagnoses of cancer (n=27), thyroid conditions (n=65) or chronic obstructive pulmonary disease (n=3) were also excluded due to the long-term effects of these conditions on skeletal muscle mass (17-19). The decision to exclude individuals who reported taking anabolic steroids was made *a priori*, however, no 5 individuals in this dataset met this criterion. The final female cohort consisted of 753 pre-menopausal females aged 18-40 years.

### Procedures and Measures

#### Demographic and Health Information and Behaviours

NHANES interviewers collected information about age, race, gender, medical history, reproductive health, dietary information, alcohol consumption and physical activity levels about all the individuals in the household. This information was gained through standardised questionnaires delivered by a trained interviewer, according to NHANES protocol, which can be found at https://www.cdc.gov/nchs/nhanes. Physical activity data was collected using the Global Physical Activity Questionnaire (GPAQ). Dietary information was collected via a 24-h recall administered by NHANES interviewers. The alcohol use variable was made from answers to questions from the home interviews. Individuals were categorised into: <12 drinks in lifetime (very infrequent), having at least one drink on one to three days per month (infrequent), having at least one drink on one to three days per week (moderate), or having at least one drink on four to seven days per week (frequent). Participants then attended a MEC for a physical examination.

#### Sex Hormone Analysis

Before arriving at the MEC, participants were randomly assigned to morning, afternoon or evening sessions. Participants in the morning sessions fasted for at least nine hours; those attending afternoon or evening sessions had no dietary restrictions. A trained phlebotomist collected blood according to NHANES protocol. Testosterone, oestrogen and SHBG were assessed via isotope dilution liquid chromatography tandem mass spectrometry (ID-LC-MS/MS). The lower limit of detection (LLOD) for testosterone, oestrogen and SHBG analysis were 0.026 nmol·L^-1^, 10.987 pmol·L^-1^ and 0.8 nmol·L^-1^, respectively, as described at https://www.cdc.gov/nchs/nhanes. The lower limit of quantification (LLOQ) was estimated by multiplying the LLOD by three.

#### Body Composition Analysis

Body composition was assessed via dual-energy x-ray absorptiometry (DEXA) during the MEC. Scans were acquired with the Hologic Discovery model A densitometers (Hologic, Inc., Bedford, Massachusetts), using software version Apex 3.2. Individuals were not eligible for a DEXA scan if their weight or height exceeded 450 pounds (204.11 kg) or 6’5” (195.58 cm), respectively. In-depth protocols for DEXA scans are located at https://www.cdc.gov/nchs/nhanes.

#### Handgrip Strength

The procedures to measure handgrip strength is described in detail at https://www.cdc.gov/nchs/nhanes. Briefly, participants squeezed a dynamometer as hard as possible with their dominant hand, in a standing position. The test was repeated on the other hand, and then twice more for each hand. Exactly sixty seconds separated attempts on the same hand. Combined grip strength was the sum of the largest reading from each hand and was used in our final analyses.

### Data Cleaning and Manipulation

Testosterone concentrations were converted to SI units (nmol·L^-1^) by dividing all data by 28.818. Any hormone data that were below the LLOQ of the ID-LC-MS/MS were removed. No individuals had testosterone readings below the LLOQ, 76 individuals had oestrogen levels below the LLOQ and no individuals had SHBG levels below the LLOQ. Implausible hormone values (pragmatically defined as values that were > mean ± eight x SD) were also coded missing. One female had testosterone levels over 19 nmol·L^-1^, one female had a SHBG value of over 1000 nmol·L^-1^, and three females had oestrogen readings over 700 pg·mL^-1^; all of these values were accordingly coded as missing.

Total physical activity levels were calculated according to NHANES and Global Physical Activity Questionnaire (GPAQ) guidelines. The average minutes per week was calculated for each discrete physical activity domain (vigorous or moderate work, transport and vigorous or moderate leisure time) and converted to metabolic equivalents (METS). Vigorous activity was classified at eight METS and moderate or transport physical activity was classified as four METS, as per NHANES protocol. The domain-specific MET scores were then summed to generate a total physical activity measurement in MET-minutes per week.

Height-adjusted lean mass, or fat free mass index (FFMI), was calculated by dividing total body lean mass (excluding bone mineral content) in kg by height squared. Fat free mass (%) was calculated by dividing total body lean mass (excluding bone mineral content) by body mass and multiplying by 100. Upper body lean mass (UBLM) was calculated by summing the lean mass for the right and left arms and dividing by height squared. This measurement was used to measure the appendicular lean mass of the upper body only. This excludes organ mass, which may be influenced factors including hydration status, and provides a more accurate depiction of muscle mass of the upper body (20). Lower body lean mass (LBLM) was calculated by summing the lean mass for the right and left legs and dividing by height squared. Free androgen index (FAI) was calculated by dividing total testosterone (nmol·L^-1^) by SHBG (nmol·L^-1^) and multiplying by 100.

All independent (total testosterone, FAI, SHBG) and dependent (FFMI, UBLM, LBLM, handgrip strength) variables were standardised by calculating the z-score for each variable (mean=0, SD=1). Standardised variables were used for subsequent analyses, allowing for estimation of the magnitude of any significant relationship.

### Statistical Analyses

All statistical analyses were performed with Stata software version 15.0 (StataCorp, College Station, TX) and accounted for the complex survey design and stratification employed by NHANES by using the appropriate sample design variables (strata and primary sampling unit). The one-day dietary weighting scheme was applied to account for oversampling of different populations and yield estimates representative of the US population, according to NHANES data analysis guidelines, found at https://www.cdc.gov/nchs/nhanes. This scheme was chosen as it relates to the smallest sampling unit, as per NHANES protocol.

Missing data were examined, and no patterns of missing data were identified. Under a missing at random assumption, multiple imputation by chained equations with predictive mean matching (using five nearest neighbours) was used to handle the missing data. Thirty imputations were used, based on 30% of participants having at least one missing data point for the study variables. All the analysis variables were used in the imputation model, with no additional auxiliary variables.

Multiple linear regression was performed to examine the relationship between testosterone (independent variable) and handgrip muscle strength and lean mass (dependent variables) separately. Initial models include both linear and quadratic terms for testosterone, to account for potential curvilinear relationships. If there was insufficient evidence of a quadratic effect (*p*-values > *0.1*), a linear model (with no quadratic effect) was tested. Covariates included in the analysis were: physical activity, ethnicity, female hormone use, oestrogen, SHBG, age, body fat percentage, alcohol consumption and the examination session to which each participant was assigned (morning, afternoon or evening; to account for the diurnal variation and the effects of fasting on testosterone concentrations and handgrip strength) (21). Dietary protein, vitamin C and D and magnesium intake were also accounted for as previous studies have shown relationships between lean mass and dietary protein, vitamin C and D and 9 magnesium intake in females (22-25). Collinearity was assessed by variance inflation factors, with a threshold of three set. No variables suggested inappropriate collinearity. All data are represented as mean ± SD. Statistical significance was set at *p<0.05*. All exposure and outcome variables were standardised to assess the effect size of any significant relationships. Coefficients <0.2 were considered ‘small’ effects, 0.2<0.5 were considered ‘medium’ effects, and 0.5<0.8 were considered ‘large’ effects.

## Results

Our final cohort consisted of 753 females aged 18-40 years, with a mean age of 30 years. Mean testosterone concentration was 1.0 nmol·L^-1^ (range 0.1 to 5.3), mean FFMI was 16.4 kg·m^-2^ (range 10.6 to 30.5) and mean combined handgrip strength was 61.7 kg (range 22.6 to 99.7). Over two-thirds (68.6%) of participants had taken female hormones throughout their life, either for contraception or other uses. The full characteristics of the study population are shown in Table 1.

**Table 1.** Weighted characteristics of included females. Values are mean ± standard deviation. n= 753

| Variable | Mean ± SD<br>($n=753$ females) | Range<br>(min-max) |
| --- | --- | --- |
| Age (years) | $29.6 \pm 6.4$ | 18-40 |
| Ethnicity (%) |  |  |
| Non-Hispanic White | 54.3 |  |
| Non-Hispanic Black | 14.6 |  |
| Non-Hispanic Asian | 7.1 |  |
| Other Non-Hispanic | 3.3 |  |
| Mexican Hispanic | 12.7 |  |
| Other Hispanic | 8.0 |  |
| Height (cm) | 162.7 ± 6.8 | 143 – 184 |
| Weight (kg) | 75.5 ± 22.0 | 37.6- 184.8 |
| BMI (kg·m<sup>2</sup>) | 28.5 ± 7.7 | 16.1 – 60.9 |
| Fat Free Mass Index (FFMI; kg·m<sup>2</sup>) | 16.4 ± 3.0 | 10.6 – 30.5 |
| Height-adjusted upper body lean mass<br>(UBLM; kg·m<sup>2</sup>) | 1.7 ± 0.4 | 1.0 – 3.3 |
| Height-adjusted lower body lean mass<br>(LBLM; kg·m<sup>2</sup>) | 5.2 ± 1.1 | 3.0 – 11.4 |
| Fat Free Mass (%) | 59.3 ± 5.6 | 45.6 – 77.3 |
| Fat percentage (%) | 37.7 ± 6.1 | 18.8 – 52.8 |
| [Testosterone] (nmol·L<sup>-1</sup>) | 1.0 ± 0.6 | 0.1 – 5.3 |
| [Oestrogen] (pg·mL<sup>-1</sup>) | 94.0 ± 79.3 | 9.0 – 51.0 |
| [SHBG] (nmol·L<sup>-1</sup>) | 81.4 ± 62.0 | 8.9 – 452.3 |
| Free Androgen Index (FAI) | 0.02 ± 0.02 | 0.001 – 0.18 |
| Combined handgrip strength (kg) | 61.7 ± 10.5 | 22.6 - 99.7 |
| Protein intake (g·day<sup>-1</sup>) | 73.0 ± 34.0 | 3.0 - 296.2 |
| Total vitamin C intake (mg·day<sup>-1</sup>) | 72.0 ± 71.4 | 0.0 - 796.3 |
| Total vitamin D intake (mcg·day<sup>-1</sup>) | 4.0 ± 5.4 | 0.0 - 62.4 |
| Total magnesium intake (mg·day<sup>-1</sup>) | 262.1 ± 137.5 | 36 - 2725 |
| Female hormone use (%) |  |  |
| No | 32.0 |  |
| Yes | 68.0 |  |
| Average total physical activity<br>(MET-min/week) | 3 347 ± 5 636 | 0 – 45 600 |
| Time of venepuncture (%) |  |  |
| Morning (fasted) | 45.3 |  |
| Afternoon | 29.8 |  |
| Evening | 25.0 |  |
| Alcohol Consumption (%) |  |  |
| <12 drinks in life | 9.6 |  |
| ≥1 drink on 1-3 days/month | 39.6 |  |
| ≥1 drink on 1-3 days/week | 40.2 |  |
| ≥1 drink on 4+ days/week | 10.6 |  |

In our cohort, there was no evidence of quadratic effects of total testosterone on total FFMI, UBLM, LBLM or handgrip strength (all p-values>0.1). There were also no significant linear effects of total testosterone on FFMI, UBLM, LBLM or handgrip strength. These results are shown in Table 2. Conversely, FAI, a measure of testosterone that is not bound to SHBG and may be therefore considered the ‘free’ portion of testosterone, was positively linearly associated with FFMI (β=0.17; *p=0.041)* and UBLM (β=0.19; *p=0.015*. There was no evidence of linear effects of FAI on LBLM or handgrip strength, nor was there evidence of quadratic effects of FAI on any variable. The linear effects of FAI on FFMI, UBLM, LBLM and handgrip strength are presented in Table 3.

**Table 2.** Linear effect of total testosterone (nmol·L^-1^) on fat free mass index (FFMI) (kg·m^-2^), upper body lean mass (UBLM) (kg·m^-2^), lower body lean mass (LBLM) (kg·m^-2^) or combined handgrip strength (kg) in 18-40 year old females (n=753)

| Variable (linear term) | β (95% CI) | *p* |
| --- | --- | --- |
| FFMI (kg·m<sup>-2</sup>) | 0.08 (-0.02, 0.18) | 0.113 |
| UBLM (kg·m<sup>-2</sup>) | 0.10 (-0.02, 0.21) | 0.092 |
| LBLM (kg·m<sup>-2</sup>) | 0.07 (-0.02, 0.17) | 0.130 |
| Combined handgrip strength (kg) | 0.03 (-0.11, 0.17) | 0.666 |

**Table 3.** Linear effect of free androgen index (FAI) on fat free mass index (FFMI) (kg·m^-2^), upper body lean mass (UBLM) (kg·m^-2^), lower body lean mass (LBLM) (kg·m^-2^) or combined handgrip strength (kg) in 18-40 year old females (n=753)

| Variable (linear term) | β (95% CI) | *p* |
| --- | --- | --- |
| FFMI (kg·m<sup>-2</sup>) | 0.17 (0.01, 0.33) | **0.041** |
| UBLM (kg·m<sup>-2</sup>) | 0.19 (0.05, 0.33) | **0.015** |
| LBLM (kg·m<sup>-2</sup>) | 0.12 (-0.04, 0.28) | 0.117 |
| Combined handgrip strength (kg) | 0.09 (-0.02, 0.21) | 0.100 |

Although it was originally included only as a covariate and not a primary outcome of this study, the observed positive relationships between FAI and FFMI and ULM led us to conduct further analyses on sex hormone binding globulin (SHBG). In females, SHBG was negatively linearly associated with FFMI (β=-0.12; *p=0.013)* and UBLM (β=-0.12; *p=0.009)* (Table 4). In addition, there was a trend *(p=0.054)* for SHBG to be linearly associated with LBLM (β=-0.08), but this did not reach statistical significance. SHBG was not associated with combined handgrip strength in a linear model, and there was no evidence of a quadratic effect of SHBG on any of the dependent variables.

**Table 4.** Linear effects of sex hormone binding globulin (SHBG) (nmol·L^-1^) on fat free mass index (FFMI) (kg·m^-2^), upper body lean mass (UBLM) (kg·m^2^), lower body lean mass (LBLM) (kg·m^2^) or combined handgrip strength (kg) in 18-40 year old females (n=753)

| Variable (linear term) | B (95% CI) | *p* |
| --- | --- | --- |
| FFMI (kg·m<sup>-2</sup>) | -0.12 (-0.21, -0.03) | **0.013** |
| UBLM (kg·m<sup>-2</sup>) | -0.12 (-0.21, -0.04) | **0.009** |
| LBLM (kg·m<sup>-2</sup>) | -0.08 (-0.17, 0.00) | 0.054 |
| Combined handgrip strength (kg) | -0.02 (-0.12, 0.08) | 0.692 |

Oestrogen may reduce muscle protein breakdown and increase muscle sensitivity to anabolic stimuli in females (12). We therefore explored the possibility of a relationship between oestrogen and FFMI, UBLM, LBLM and handgrip strength. However, there was no evidence for linear of quadratic effects of oestrogen on FFMI, UBLM, LBLM or handgrip strength (all p-values>0.1).

## Discussion

Despite the essential role of skeletal muscle for whole-body movement and metabolism, our understanding of the role of testosterone in muscle mass and strength has been mostly gained from male-only cohorts, warranting female-specific investigations. In line with our hypothesis,data from pre-menopausal women (aged 18-40 years) of the NHANES dataset indicate that total testosterone is not associated with FFMI or handgrip strength in females. To our knowledge, this relationship has not been tested in a large, representative population of healthy, pre-menopausal women before.

Previous, smaller studies have suggested that total testosterone is not related to lean mass in young females. Total testosterone was not associated with lean mass in healthy 18-40 year old females (n=185) (26). Furthermore, in lean women (BMI approximately 22 kg·m^-2^) aged 17-21 years with ovarian hyperandrogenism, defined by amenorrhea or oligomenorrhea and/or hirsutism, lean mass was significantly reduced when compared to healthy controls (n=22/group) (27). This finding was replicated in a small cohort of lean (BMI <25 kg·m^-2^) women aged 18-30 suffering from polycystic ovary syndrome (PCOS; n=10/group) (28). PCOS is the most common cause of hyperandrogenism in women and affects as much as 4-10% of reproductive-aged women (29) and 20-37% of elite female athletes (30). PCOS participants had significantly lower lean mass than their weight and BMI-matched, healthy counterparts, despite having higher testosterone concentrations and similar levels of SHBG, oestradiol, follicle-stimulating hormone and luteinising hormone (n=10/group) (28). In contrast, a small study observed a positive association between FFMI and total testosterone in young women with PCOS (n=48) (31), where only subjects with a FFMI above 14 kg·m^-2^ displayed significantly higher total testosterone (31). Of importance, this model was not adjusted for potential confounders such as SHBG, physical activity or diet, all of which may affect lean mass (22-26).

*In vivo*, testosterone exists either ‘free’ and unbound, or bound to proteins, such as albumin or SHBG (32). It was historically assumed that only the ‘free’ form of testosterone was able to exert its effects on cells. There is however evidence suggesting that protein-bound steroids can also activate anabolic pathways, such as the Akt/mTOR pathway in rat myocytes *in vitro* (33).

Protein-bound sex steroids can also be internalised by cells via endocyctosis in female and male mice, suggesting that they may be biologically active (34). In population studies, the free proportion of androgens is more commonly related to muscle mass and strength in males and females than the total concentration (35). In line with the existing literature, our results provided evidence for a positive association between FAI, defined as the ratio between total testosterone and SHBG levels, and FFMI and UBLM. FAI has been previously positively associated with lean mass in 18-40 year old females (n=95 PCOS patients, 90 healthy controls) (26); a relationship that dissipated when the model was adjusted for insulin (26). Insulin may mediate SHBG levels by decreasing hepatic SHBG production (36), thereby influencing the FAI and its association with lean mass in females. Women suffering from PCOS with high insulin and IGF-1, another anabolic hormone, consistently display low SHBG concentrations (37-40). One limitation of our model is that it was not adjusted for insulin or IGF-1, as insulin was only measured in females who took part in the morning session of the MEC (constituting less than half of our cohort) and IGF-1 was not measured by NHANES at all; a constraint to bear in mind when interpreting our findings. In line with these results, we also found evidence of a negative relationship between SHBG and FFMI and UBLM. Taken together, our results and others (26) suggest that, in young women either healthy or suffering from PCOS (n=95), the regulation of lean mass in pre-menopausal women may be more strongly mediated by SHBG, via its capacity to bind to testosterone, than by total testosterone itself. Evidence however suggests that SHBG may be more than simply a transport protein (34, 41). Indeed, a SHBG receptor exists on the membrane of rat skeletal muscle (41). Upon ligand binding, this activates cAMP as a secondary messenger to mediate the actions of androgens on their target cells (41, 42). SHBG might therefore also mediate the actions of steroids *in vivo* and provides interesting opportunities for future research. While FAI and SHBG were associated with FFMI and UBLM, the small coefficients (β-values <0.2) should however be kept in mind as they suggest that only a minor proportion of the variability of FFMI and UBLM can be explained by steroid concentrations. This highlights the complexity of human physiology, where a myriad of different internal and external factors influence muscle health.

Interestingly, our results indicated an association between FAI and SHBG and UBLM, but not LBLM. In males, AR protein expression is greater in upper body musculature, when compared to lower body (43). It is unknown whether females exhibit this same differential expression, which may suggest that upper body appendicular musculature is more sensitive to androgens than lower body musculature. Indeed, previous research showed a negative relationship between testosterone and trunk:leg muscle ratio and a positive relationship between testosterone and trunk:leg fat ratio in young women (aged 20-38 years) with PCOS (n=67) (44). This suggests that higher levels of free testosterone promote an ‘android’ body shape, with more fat and muscle deposition in the upper compartments of the body.

In line with our hypothesis, there was no evidence of relationships between strength and total testosterone, SHBG or FAI. This is an important set of findings, as the functional capacity of a muscle, measured by muscle strength, is arguably more important than muscle size in both athletic and every-day circumstances. Females can exhibit significant increases in muscular strength with training without a concurrent increase in muscle mass (5). This suggests that neural adaptations are primary drivers of strength gains in women, rather than muscle hypertrophy (5). Muscle strength may therefore be a more important determinant of athletic performance than muscle mass, as previously shown in older adults (45). Our results indicating no associations between markers of androgenicity and handgrip strength in females suggest that, while FAI and SHBG are related to FFMI, this may not necessarily translate to muscle strength and athletic ability. It should however be noted that, despite being a commonly used and robust measure of overall muscular strength (46), handgrip strength is not a perfect proxy for whole-body muscle strength (47) and this should be accounted for when interpreting the results.

Recently, endogenous testosterone levels have been used as an eligibility criterion for specific athletic events in female sports (48). Hyperandrogenic athletes (defined as females with testosterone levels over five nmol·L^-1^ (49)) are banned from competing in specific athletic running events on the bases of their naturally-occurring testosterone levels and the assumption of a direct association with athletic performance. Our data suggest that FAI, but not total testosterone, and SHBG are associated with FFMI. However, neither SHBG nor FAI were associated with LBLM. This is an interesting finding in the context of the eligibility of hyperandrogenic athletes for running events. Indeed, it may be argued that lean mass and strength endurance of the lower body are more relevant measures than total body lean mass or upper body lean mass, when considering which events these regulations affect, including running events ranging 400 m to 1500 m (49). Taken together, our data and others (26-28) suggest that more evidence is needed to validate such a relationship in both healthy and hyperandrogenic women.

When compared to the previous literature, the current study includes an unprecedentedly large sample size that is representative of the American population. Furthermore, the models used in this study have been adjusted for a number of covariates that can influence lean mass in females, which constitutes a strength of our analyses when compared to previous research. Using FFMI and FAI in our models also provides a more physiologically relevant picture of the relationship between androgens, muscle mass and muscle strength in females. FFMI accounts for the height of individuals, as opposed to lean mass as an absolute measure. Further, our study divided lean mass into upper and lower body compartments. This offers new insights into the differential effects of testosterone on different body compartments and suggests that upper body appendicular musculature may be more sensitive to androgens than lower body musculature in females.

Limitations include the cross-sectional, observational data, which prevents inferring causal relationships. In addition, NHANES does not include growth hormone (GH) or insulin-like growth factor-1 (IGF-1) measures or complete insulin data. These anabolic hormones may play a role in the regulation of lean mass or muscle strength in females and should be accounted for as covariates when possible.

In conclusion, our data indicate that total testosterone is not related to FFMI, UBLM, LBLM or handgrip strength in pre-menopausal females, suggesting that it is not a direct determinant of lean mass or muscle strength in this population. Our findings also indicate a positive relationship between FAI and lean mass, and a negative relationship between SHBG and lean mass. When compared to the total pool, ‘free’ testosterone concentrations may therefore be more highly associated with lean mass in females. In addition, androgens may also be more highly associated with upper body lean mass compared to lower body lean mass in females. Further, longitudinal or interventional research is warranted to better understand these relationships.

## Data Availability

This manuscript uses freely available data from the National Health and Nutrition Examination Survey (NHANES) from the 2013-2104 cycle.

https://www.cdc.gov/nchs/nhanes/index.htm

## Acknowledgements

Funding: SA is supported by the Australian Government Research Training Program (RTP). No other funding was received as part of this project.

